# Potential contribution of climate conditions on COVID-19 pandemic transmission over West and North African countries

**DOI:** 10.1101/2021.01.21.21250231

**Authors:** Ibrahima Diouf, Souleymane Sy, Habib Senghor, Papa Fall, Diarra Diouf, Moussa Diakhaté, Wassila M. Thiaw, Amadou T. Gaye

## Abstract

The COVID-19 disease, caused by the Severe Acute Respiratory Syndrome Coronavirus 2 (SARS-CoV-2), is a very contagious disease that has killed many people around the world. According to the World Health Organization (WHO) data, the spread of the disease appears to be slower in Africa. Although a number of studies have been published on the relationship between meteorological parameters and COVID-19 transmission, the effects of climate conditions on COVID-19 remain largely unexplored and without consensus following the main research finding over Africa (often based on a single country or city). Here, using available epidemiological data over 275 days (i.e., from March 1 to November 30, 2020) taken from the European Center for Disease Prevention and Control of the European Union database and daily data of surface air temperature and humidity from the National Center for Environmental Prediction (NCEP), this paper investigates the potential contributions of climate conditions on COVID-19 transmission over 16 countries selected from three bioclimatic regions of Africa (i.e., Sahel, Maghreb and Gulf of Guinea). On average, our main findings highlight statistically significant inverse correlations between COVID-19 cases and temperature over the Maghreb and the Gulf of Guinea regions, whereas positive correlations are found in the Sahel, especially over the central part including Niger and Mali. Correlations with specific humidity and water vapor parameters display significant and positive values over the Sahelian and the Gulf of Guinean countries and negative values over the Maghreb countries. In other word, results imply that the COVID-19 pandemic transmission is influenced differently across the three bioclimatic regions: i) cold and dry environmental conditions over the Maghreb; ii) warm and humid conditions over the Sahel iii) cold and humid conditions over the Gulf of Guinea. These findings could be useful for decision-makers who plan public health and control measures in affected African countries and would have substantial implications for directing respiratory disease surveillance activities.

## Introduction

The coronaviruses constitute a family of various viruses affecting both humans and animals. In late December 2019, the World Health Organization (WHO) was informed about an epidemic of “pneumonia of unknown cause” detected in the city of Wuhan (Hubei Province, China), the seventh-largest city in China, with 11 million inhabitants [1,2]. The first reported infected individuals were associated to the Wuhan seafood market in southern China. The virus causing the epidemic was quickly determined to be a novel coronavirus linked to the Middle Eastern Respiratory Syndrome Coronavirus (MERS-CoV) and to the Severe Acute Respiratory Syndrome Coronavirus 2 (SARS-CoV-2) [3]. Infected travelers, mainly by aircraft, are known to be responsible for the introduction of the virus outside Wuhan [4]. The novel coronavirus so-called 2019-nCoV in the early parts of the epidemic has spread rapidly to multiple countries and has been declared on March 11, 2020, as a pandemic by the WHO. The 2019-nCoV terminology changed later and become Coronavirus Disease 2019 (COVID-19).

Researchers who have been working on this epidemic believe that this coronavirus probably originates from bats [5,6]. This hypothesis is based on the SARS epidemic in 2003 that emerged in southern China after being transmitted from the bat, its “natural reservoir", to humans [7]. Indeed, the COVID-19 disease shares 80% of genetic similarity [7]. The infection caused a wide range of symptoms and showed different degrees of severity with different fatality rates that are country dependent. Symptoms may appear 2-14 days after exposure to the virus [8]. People with these symptoms or combinations of symptoms may have cough and shortness of breath or difficulty breathing. Transmission can take place by air, in contact with secretions or contaminated objects. The mortality rate is found to be relatively lower compared to SARS-Cov which was estimated around 15% [8]. The most sensitive people are the elderly, those suffering from respiratory and immunosuppressed pathologies.

One of the central questions facing the scientific community is how climate conditions contribute to COVID-19 transmission. Chan et al.,[9] investigations on the past SARS coronavirus outbreaks have put in evidence that the viability of the virus was lost at high temperatures (e.g., 38°C) and high humidity (e.g., > 95%) and this explains the lower number of cases in Asian countries including Malaysia due to higher temperature and humidity. The first studies from China have shown that the contagiousness of the COVID-19 is higher in the cities in the north with a lower humidity value and lower temperatures than those over the south-eastern coasts with warm and humid conditions [10,11]. Meteorological variables such as air temperature, humidity, and other parameters including solar radiation have been also found to act differently concerning coronavirus survival [12]. Although several studies have supported the hypothesis that climate conditions may affect the COVID-19 pandemic transmission as happens with other viruses such as influenza [13,14], respiratory syncytial virus [15,16], considerable disagreement is evident among the different investigations (often based on different country or city); most of them have concluded that the COVID-19 transmission prefers cold and dry weather conditions -i.e. temperature and humidity related variables are negatively correlated to the virus transmission [12,17–22] and [23] (for a review) -, while others studies have reported contradictory results showing that temperature and/or humidity variables may positively associated, or not be associated with the COVID-19 expansion [24–28]. The majority of the above-mentioned papers were focused on Asian and European countries (i.e., mid-latitude regions). Few studies have investigated the effects of weather conditions on the incidence of COVID-19 pandemic transmission over Africa and the results have even less consensus (often based on a single country or city) [29–31].

In addition, it is worth pointing out that since the first cases of coronavirus appeared in Africa (early March 2020), the spread of the disease seems to be progressing more slowly there than elsewhere. The five most affected countries at this time are South Africa, Algeria, Morocco, Egypt, and Cameroon, with a very slow infection rate compared to the mid-latitudes or south America countries (see, Fig 1). Nevertheless, it is still not fully understood how climate conditions contribute to COVID-19 transmission in Africa.

**Fig 1.**
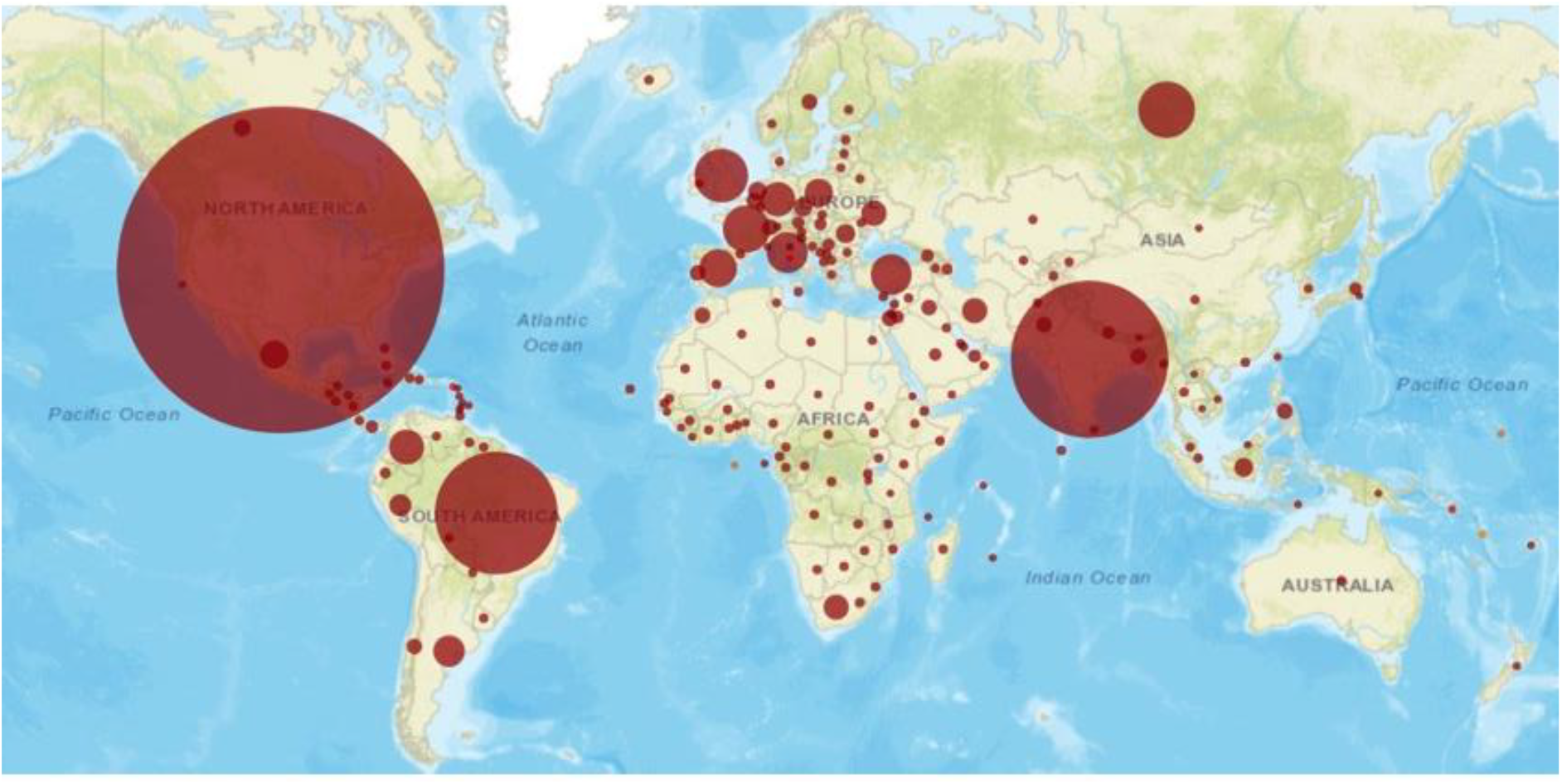
COVID-19 cumulative infection cases by country. The map of COVID-19 pandemic transmission shows the most affected countries at this time are: US, India, Brazil, UK and Russia with a very slow infection rate in African countries. Source: https://infographics.channelnewsasia.com/covid-19/map.html

The outlined scenario shows that further studies and reviews are needed to extend the analysis available in literature and to quantify the possible relationship between climate conditions and coronavirus transmission in Africa. To address these issues, here, using available epidemiological data over 275 days (1 March -November 30, 2020) extracted from the European Center for Disease Prevention and Control of the European Union (https://www.ecdc.europa.eu/en/publications-data/download-todays-data-geographic-distribution-covid-19-cases-worldwide) and daily data of surface air temperature and humidity from the National Center for Environmental Prediction (NCEP), the present study aimed to investigate the potential contributions of climate conditions on COVID-19 pandemic transmission in 16 highly populated West and North African countries divided into three bioclimatic regions (i.e., Maghreb, Sahel and Gulf of Guinea) [32].

The present work is organized as follows: Section 2 outlines the data used and the statistical methods applied. Section 3 investigates the spatial distribution of COVID-19 confirmed cases throughout countries in North and West Africa and their potential relationship with climate conditions. Finally, a summary and discussion of the main findings are provided in Section 4.

## Data and Methods

### Surveillance COVID-19 data

The clinical data corresponds to the number of COVID-19 cases over 16 West and North African countries recorded by the European Center for Disease Prevention and Control of the European Union, and all age-groups were screened (https://www.ecdc.europa.eu/en/publications-data/download-todays-data-geographic-distribution-covid-19-cases-worldwide). The countries were selected based on their high population density and the regularity of the reported daily positive cases. The selected countries include the Maghreb countries (Algeria, Egypt, Libya, Morocco and Tunisia), the Sahelian countries (Burkina-Faso, Mali, Niger and Senegal) and Gulf of Guinea countries (Ivory Coast, Ghana, Guinea Conakry, Liberia, Nigeria, Sierra Leone and Togo). Note, the number of observed COVID-19 cases is available for the selected countries from March 1 to November 30, 2020 and datasets are obtained at country level. These cases are clinically confirmed by the Polymerase Chain Reaction (PCR) tests, as recommended by the WHO.

### Climate dataset

Due to the lack of recent continuous observed weather station datasets over the selected countries, reanalysis climate datasets are used. Reanalysis is a systematic approach to produce datasets for climate monitoring and research [33]. We use daily data of surface air temperature, relative humidity, and specific humidity from NCEP datasets provided by the NOAA [33] with 2.5°× 2.5° horizontal resolution over the North and West Africa. Besides those variables, the Water Vapor variable (WV; g/kg) is estimated following [11,14,34–40] methodology (see equation 1), where RH represents the relative humidity, T the air temperature, and P the atmospheric pressure. Note the climate variables for the period from February 15 to November 30, 2020 (i.e., 15 days before the first reported COVID-19 cases) are selected in the analysis to take into consideration the virus incubation period not considered in many investigations over Africa [29,30,41]. In other term, because of 97.5% of the infected people develop symptoms after 11 days of incubation [42], climate variables are 15-days back time-shifted. This imply that the daily number of positive COVID-19 cases from 1 March 2020 to 30 November 2020 are the result of infections that happened from 15 February 2020 to 15 November 2020. This approach is inspired to [43].

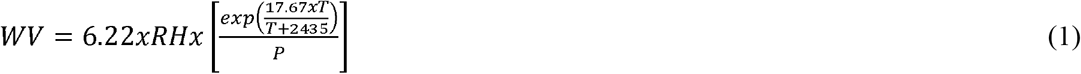

Africa through the northern part is mainly characterized by different types of climates [44], as we mentioned above, the selected countries were divided into three contrasted bio-climate zones based on a classification that closely fits with Köppen-Geiger bio-climatic zones [32,45]:

i. the Maghreb (North Africa) countries are mainly characterized by winter rains and summer drought [46]. The rainfall regime is variable but predominantly bimodal in north part, with fall and spring peaks. The rainy seasons correspond with the short days–cool temperature period (from November to April). This category includes hyper-humid to hyper-arid bioclimates, with mean annual rainfalls of 2,330 mm over the North East of Algeria, and 1,530 mm over the North West of Tunisia [32] decreasing to virtually zero in the central–eastern Sahara. Mean annual temperature may vary from less than 10°C in the highlands of Algeria and Morocco to 25 °C in the north and central Sahara [32];
ii. the Sahel region represents a transition zone between the Saharan desert and the wet climate of tropical Africa. The Sahelian countries have a tropical semi-arid climate which is typically hot, sunny, dry and somewhat windy. The bio-climate is characterized by a monomodal (unimodal) type of rainfall distribution pattern, i.e., there is only one annual peak in the rainy season, lagged with the summer solstice by a 1–2-month time lag [32,45];
iii. the Gulf of Guinea with equatorial bio-climate, always humid, are characterized by a bimodal rainfall distribution pattern (i.e., there are two peaks in the rainy season following the seasonal position Intertropical Convergence Zone (ITCZ)), although some particular climatic characteristics can be found in countries like Nigeria with hot-dry, hot-humid, temperate-dry, temperate-humid, and temperate-dry with a cool climate [47].

### Statistical Analysis

Two non-linear robust and non-parametric rank correlation tests were implemented to analyze the potential effects of the climate conditions on the COVID-19 pandemic transmission and to discuss their statistical significance:

i. the Kendall rank correlation non-parametric test τ is showed in Equation 2, where concor represents the number of concordant pairs, while discor represents the discordant pairs, and n is the number of pairs [48].

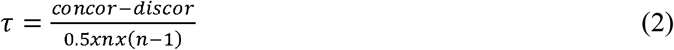
ii. The Spearman rank correlation non-parametric test r_s_ is described in Equation 3 below, where d_i_ represents the difference between the ranks of two parameters and n the number of alternatives [48].

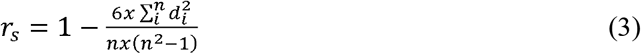

Note more descriptions of the statistical methods used can be found in [42,47]. Nevertheless, it is worth pointing out that values of τ and r_s_ equal to +1 and -1 implying a perfect positive and negative association/correlation, respectively. The main reason for using the non-parametric Kendall and Spearman statistical test is that it tends to be more powerful and better suited for non-normally distributed variables compared with parametric statistical tests such as the Pearson test [48]. As the distribution of COVID-19 cases data and climate variables are not necessarily Gaussian, the non-parametric Kendall and Spearman statistical tests might be more appropriate for testing the null hypothesis of the association/correlation between the two given variables [49,50].

In addition, the correlation coefficients are often computed between two time series (e.g., two time series representing, respectively, the number of daily COVID-19 cases and daily average temperatures or humidity) [29–31], without taking into account for the possible presence of cycles or temporal trends in the data, which can strongly affect the estimated correlation values and yield artefactual associations. In the present paper, the daily cases of COVID-19 are fitted with the robust and non-parametric Median-Based Linear Model (MBLM) defined in [49]. As it can be easily observed (see section next section) that the COVID-19 cases data does not follow a Gaussian distribution, however as discussed above, the non-parametric MBLM model might be more powerful and better suited compared with parametric linear models [49].

## Results and Discussion

### Spatio-temporal variability of COVID-19 cases

Fig 2 shows the patterns of the total number of COVID-19 confirmed cases between March 1, to November 30, 2020 over all the considered countries. It becomes obvious that the COVID-19 pandemic transmission varies in different regions during the study time period. The highest outbreak ranging up to more than ∼300000 cases is located in the Maghreb countries mainly including Morocco and Egypt. The number of COVID-19 cases ranges between 5000 to 10000 cases in the Sahelian countries and 50000 to 200000 cases in the Gulf of Guinea countries. The lowest number of cases is observed over in the central of the Sahel.

**Fig 2.**
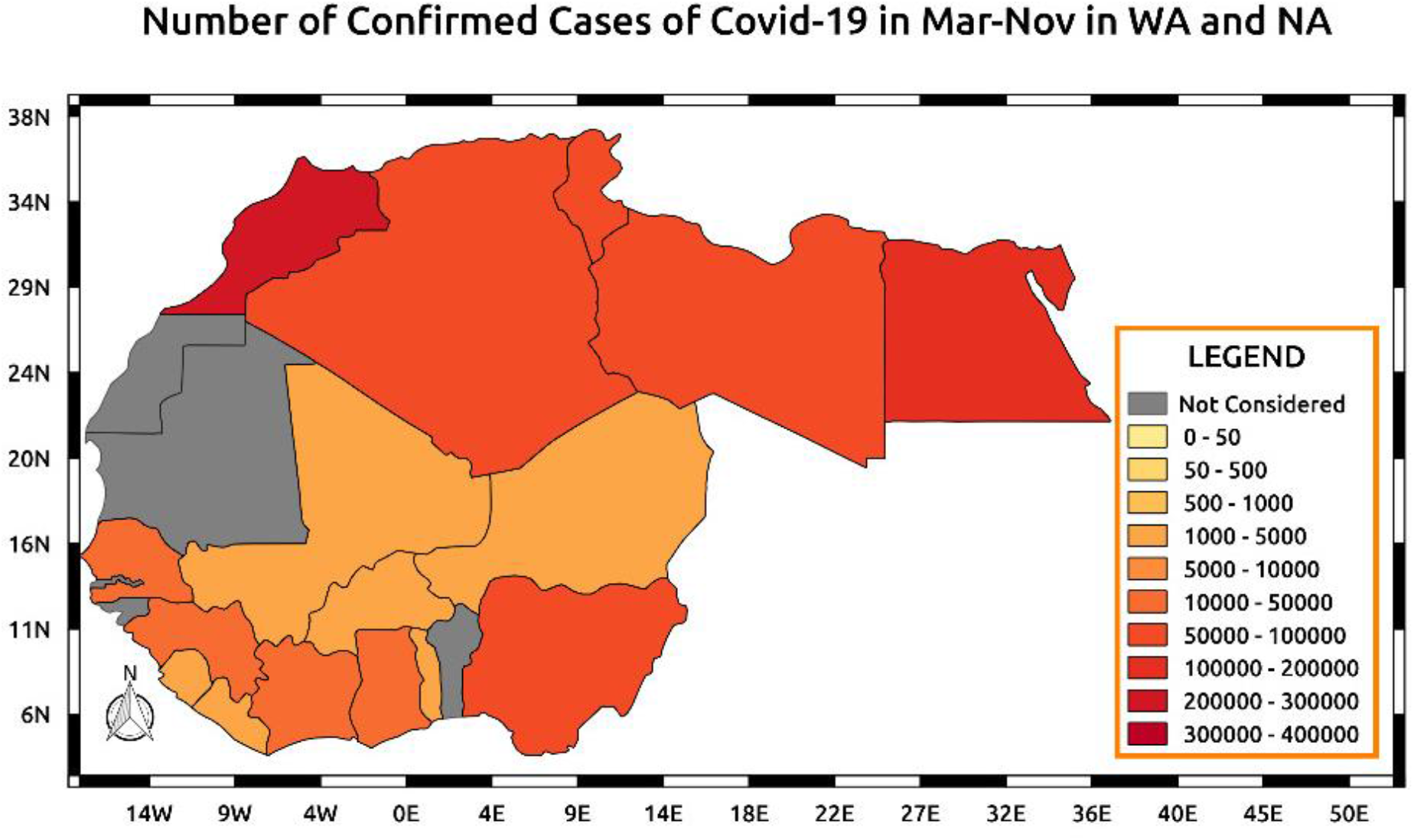
Spatial patterns of the total number of COVID-19 confirmed cases between March 1, to November 30, 2020 over all the considered countries. The countries were selected based on their high population density and the regularity of the reported daily positive cases.

Fig 3 further shows that more occurrences of COVID-19 cases are observed over the northern part of Africa in Maghreb countries compared to the Sahel and the Gulf of Guinea. Fig 3A, with y-axis is capped at ∼400000 COVID-19 cases for Morocco and 80000 for the other northern countries, shows a rapid spread of COVID-19 cases in the Maghreb with the largest trend for Morocco and Egypt. Until the end of July, the evolution of confirmed COVID-19 cases in Tunisia, and Libya was very slow, but a rapid increase is also noted since August. For the Maghreb, it is only in Libya that very few COVID-19 cases are recorded, Tunisia does not exceed 1000 cases in three months and a half, while the maximum number of cases are developed in the Maghreb countries like Egypt, Algeria, and Morocco. In Fig 3B related to the Sahel; the largest number of COVID-19 cases are observed in Senegal with a rapid expansion since the pandemic start. A stationary situation was observed since mid-September. A more homogeneous distribution is found between the rest of Sahelian countries (Fig 3B) where the y-axis could be capped at 3000 COVID-19 cases instead of 15000 cases like in Senegal. In the Gulf of Guinea (Fig 3C) with COVID-19 approaching 50000 in Ghana and 70000 cases in Nigeria after night months of the pandemic. Some small West African countries such as Sierra Leone, Liberia, and Togo have cumulated COVID-19 cases less than 1000 cases, while some countries such as Ivory Coast and Guinea Conakry exhibit an intermediate situation. For the 3 subzones, the first two months of the time period are marked by overall flat curves of the cumulated COVID-19 cases.

**Fig 3.**
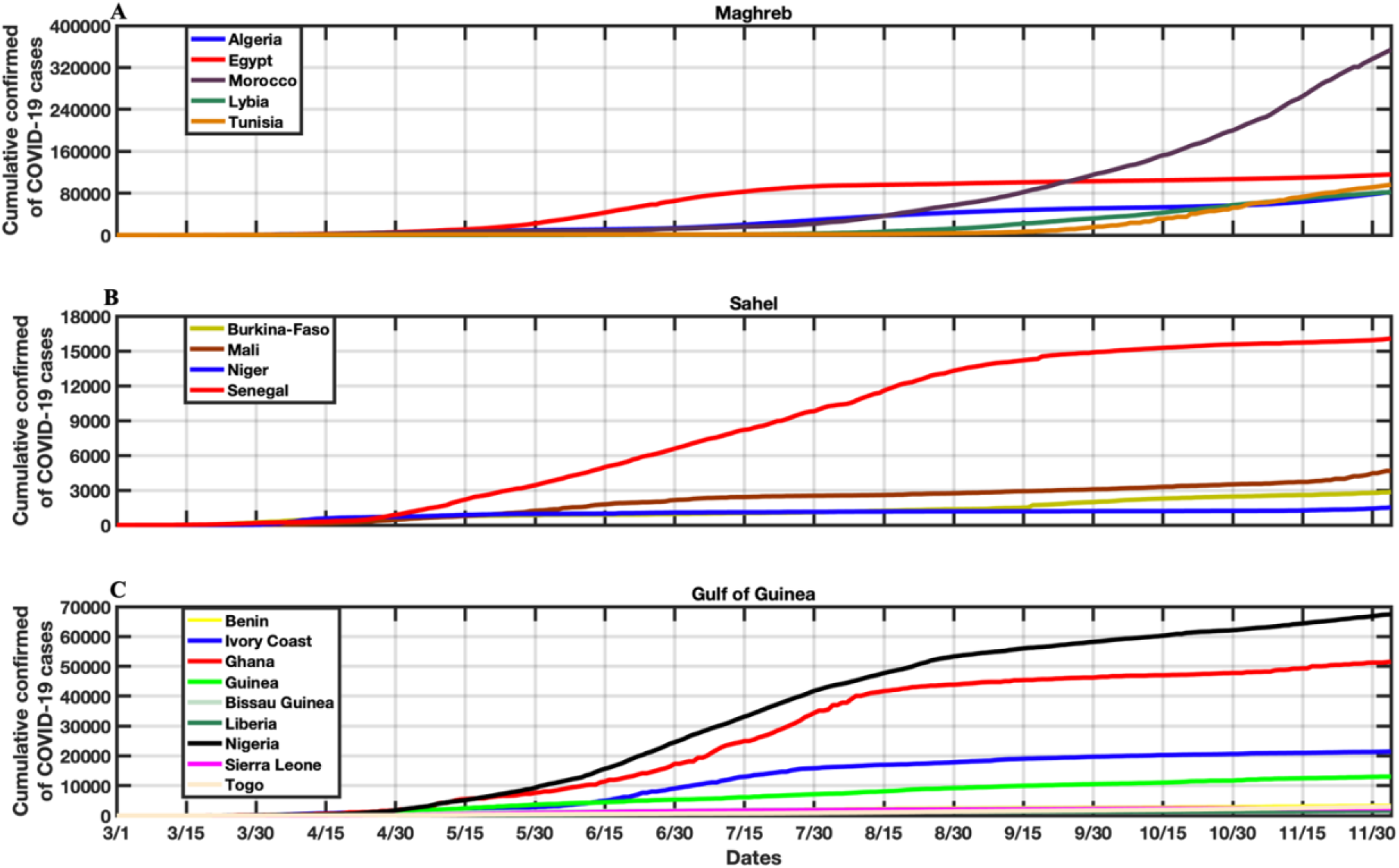
Growth curve of the cumulative confirmed COVID-19 cases over A) the Maghreb countries, B) over the Sahel countries and C) the Gulf of Guinea countries (between March 1 to November 30, 2020). Due to the large difference in the amplitudes between the three different African bio-climatic zones, the y-axis is capped at 400000 cases for Maghreb, 18000 cases for Sahel and 70000 cases for Gulf of Guinea.

In Fig 4, we compare the fitted COVID-19 cases with MBLM model to the daily observed cases for each of the three bio-climatic regions. The residuals with respect to the reported data (i.e., the differences between the MBLM fitted curve and the observed curve) are considered. Considering the residuals, makes the analysis independent on the period of lockdown or restriction, being the correlation analysis strongly dependent on the considered time period. The MBLM model accounts for the natural trend of viral epidemies and the effect of the lockdown on it. This approach is also followed in [43]. Thus, the residual analysis should preserve from spurious correlations between the above-mentioned effects and the parameters under analysis. The Spearman and Kendall rank correlations have been then computed between the residuals of COVID-19 cases with respect to the MBLM model, extrapolated from the data trend, and the detrended anomaly of the meteorological parameters with 15 days back time-shifted to take into account the virus incubation period as discussed above.

**Fig 4.**
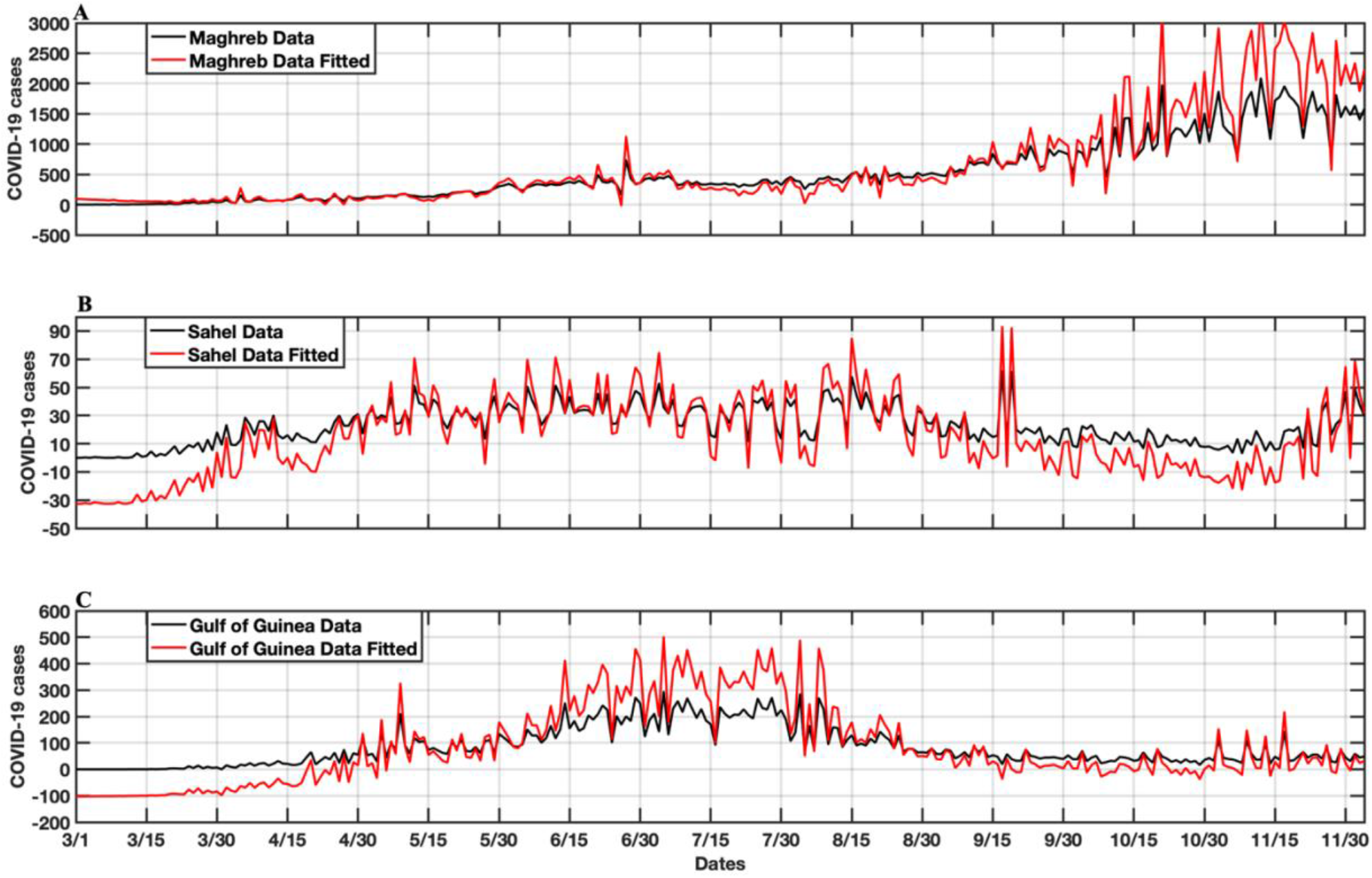
COVID-19 confirmed cases fitted by a MBLM model [49], extrapolated from the observed COVID-19 data over the A) Maghreb, B) Sahel, and C) Gulf of Guinea regions. The residuals (i.e., the differences between the MBLM fitted curve and the observed curve) (Figs. 6A, 6C and 6E) are used to investigate the correlation with the detrended anomaly of the meteorological variables.

Although a number of studies have considered the relationship between meteorological parameters and positive COVID-19 cases, it worth pointing out that the daily new positives variable is highly correlated to the number of performed test, i.e., the more the test performed, the more positives COVID-19 cases are found. In addition, delays in processing tests and false positives/negatives, being not uncommon and frequently reported, are factors that might introduce a bias in the analysis. For this reason, daily spikes in cases, without considering the incubation period, can be totally uncorrelated with the climate variables as discussed in [40].

### Spatio-temporal variability of climate parameters

Fig 5 shows the seasonal patterns of climate parameters including temperature, specific humidity and water vapor variables through the North and West Africa. A strong seasonal temperature regime (with high-temperature values during JJA and lower temperature values during September-November) associated with a slight seasonal humidity regime is observed. Comparing the seasonal patterns of climate parameters across the three seasons (i.e., during MAM, JJA, and SON), the highest values of temperature are located over the North part of the Sahel during summer (Fig 5A), while the lowest values are found over the Gulf of Guinea countries (Fig 5B). The low values of humidity and water vapor are found in the Northern area up to the central part of the Sahel (Figs 5D, E and F). In the Gulf of Guinea part, the temperature decreased from March to August, while the specific humidity and water vapor increased (Figs 5B-5H) mainly due to the fact that the Gulf of Guinea area usually received substantial rains sustaining associated with high humidity and lower temperatures. Whilst over Sahelian countries, low values of humidity are observed compared to the Gulf of Guinea part, with a wetter period in summer associated with the northward shift of the main tropical rainfall band. Over the Maghreb countries, a well-marked cool period is observed during the spring and is associated with a slight decreasing of humidity and water vapor content. These seasonal climate conditions may differently contribute to enhancing/reducing COVID-19 pandemic transmission across the different considered regions.

**Fig 5.**
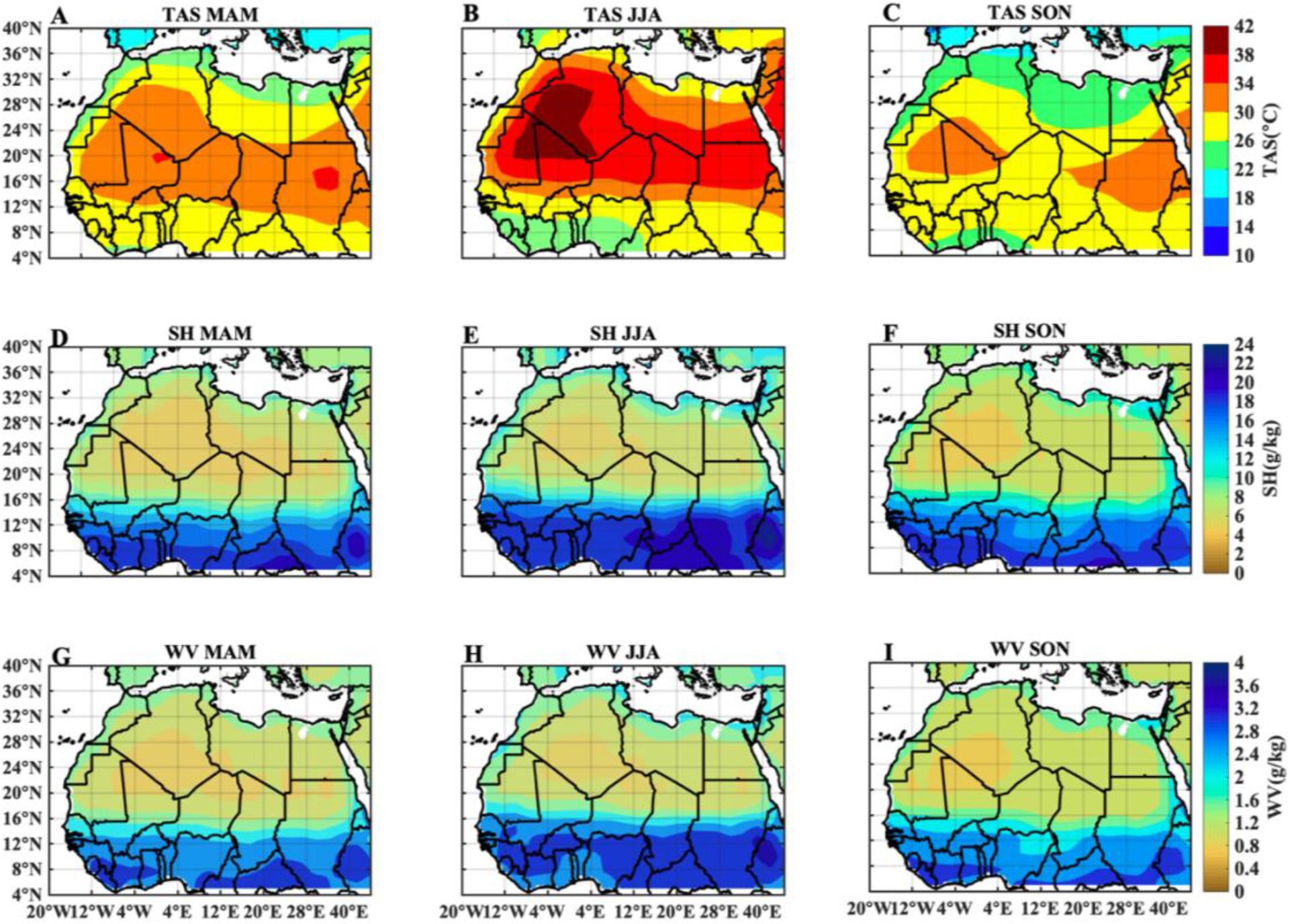
Seasonal spatial patterns of climate parameters over the North and West African countries (20°W-40E; 4°N-40°N). A) surface air temperature in MAM, B) in JJA and C) in SON; D) specific humidity in MAM, E) in JJA and F) in SON; G) water vapor in MAM, H) in JJA and I) in SON

In Fig 6, the daily COVID-19 cases anomaly (residuals) with respect to the climate parameters over the three bio-climatic zones are shown. In the Maghreb, the positive anomalies of the daily COVID-19 cases are clearly observed when the climatic parameters (i.e., temperature, humidity and water vapor) have simultaneously a downward trend as at the beginning and towards the end of the time period (Figs 6A and B). However, for the Sahel (Figs 6C and D) and the Gulf of Guinea (Figs 6E and F), the positive anomalies of COVID-19 cases are found during the summer, when these regions experience a high value of humidity and water vapor.

**Fig 6.**
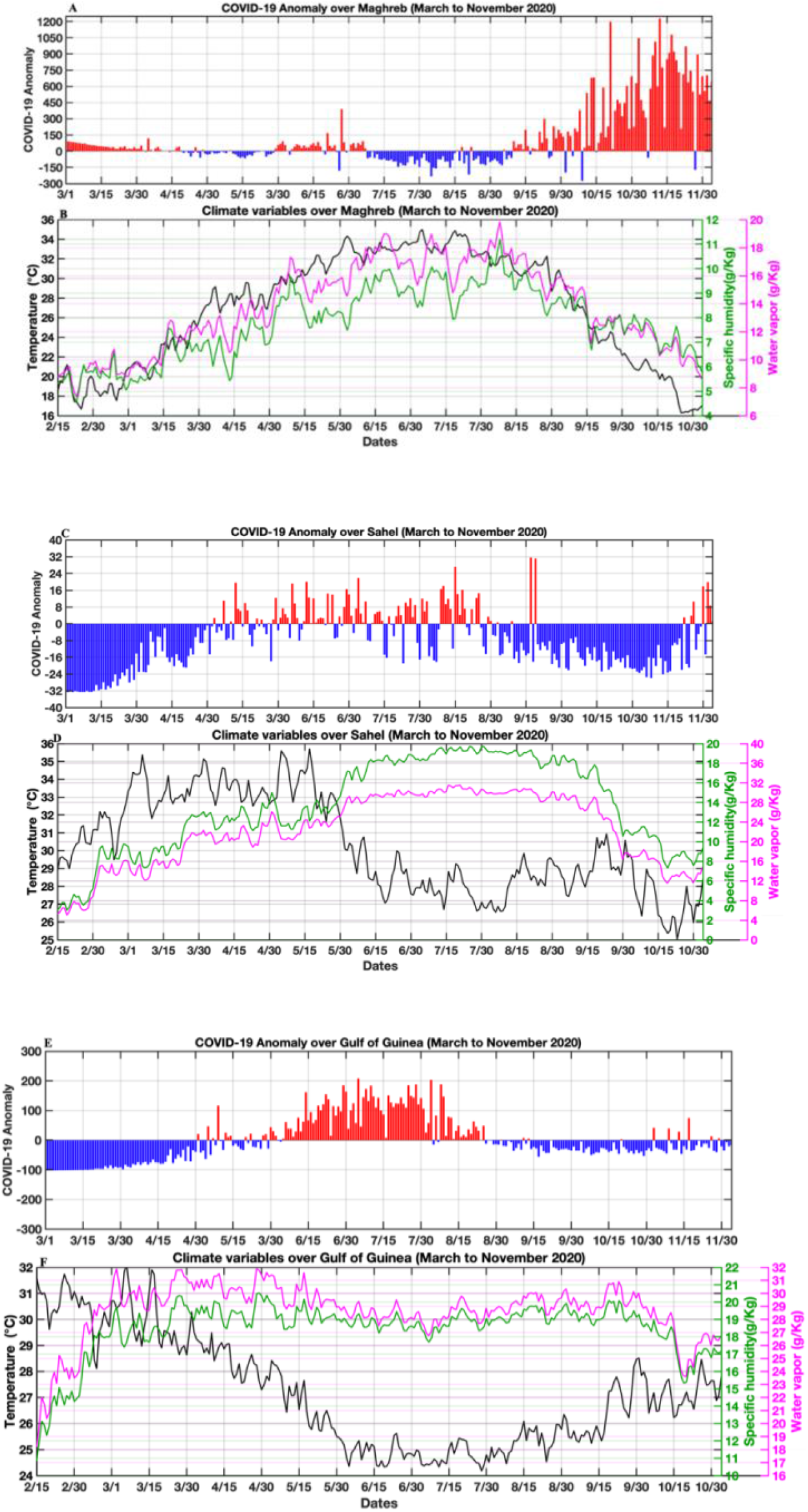
COVID-19 daily case anomaly (residual) and climates parameters for the period between March 1 and November 30, 2020. A and B) for Maghreb, C and D) for Sahel and E and F) for Gulf of Guinea.

In S1 Table, the daily variation of the meteorological parameters is also shown, together with a statistical analysis. It can be noticed that the mean temperature in Maghreb countries was 25.86±6.24°C (mean± Standard deviation) with low humidity of around 7.51±1.60 (g/kg) and water vapor around 13.18±3.40 (g/kg). Over the Sahel, the mean temperature was estimated 29.96□2.76 °C with high amount humidity (12.65±5.23; g/kg) and water vapor (20.72±8.37 g/kg) two times larger compared to the Maghreb countries. In the Gulf of Guinea, the mean temperature was also estimated 27.39±2.16 °C with high amount of humidity (17.67±2.89; g/kg) and water vapor (27.50±4.21; g/kg) during the considered period (i.e., from March 1, to November 30, 2020).

### Relationship between COVID-19 and climate parameters

The contribution of the meteorological variables (temperature, humidity and water vapor) on the COVID-19 pandemic transmission in Africa were investigated and correlation coefficient values are estimated using the robust and non-parametric Kendall (τ) (Fig 7) and Spearman (S1 Fig in supplementary material) rank tests. In Fig 7 (and in S1 Fig), the statistically significant correlations with 99% confidence intervals, that is, correlations that passed the Kendall/ Spearman rank tests at level (i.e., p < 0.01; 99% C.I), are reported in green dots and those that passed the Kendall/Spearman rank tests at 0.05 level (i.e., p < 0.05; 95% C.I), are reported in black dots. As described above, the correlation is investigated against the residual of COVID19 cases with a time-shift of 15 days, i.e., meteorological data from 15 February 2020 to 15 November 2020 and daily number of positive COVID-19 cases from 1 March 2020 to 30 November 2020 to take into consideration the incubation period. Note, we have chosen to show the correlation values calculated with Kendall test in Fig 7, because it is more insensitive to error and discrepancies in data, with p-values more accurate than Spearman’s rho test [48].

**Fig 7.**
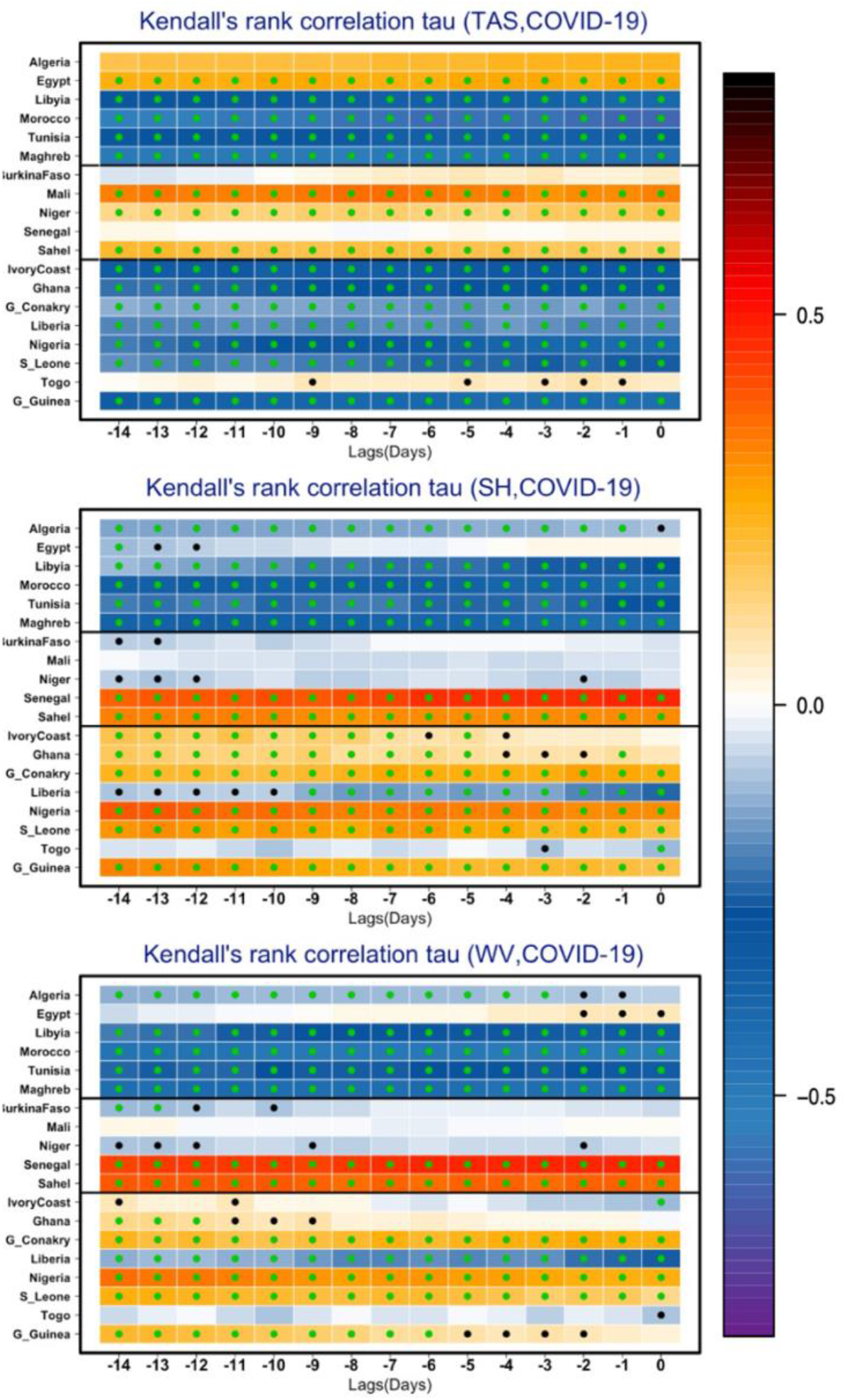
Kendall non-linear rank correlation (τ) test between the detrended anomaly of the meteorological variables and the residuals of COVID-19 cases over the selected Northern and Western African countries, as well as area averages over the three bioclimatic regions: the Maghreb, the Sahel and Gulf of Guinea. Significant correlations with 99% confidence intervals, that is, correlation that passed the Kendall rank tests at 0.01 level (i.e., p < 0.01; 99% C.I), are reported in green dots, while correlations that passed the Kendall rank tests at 0.05 level (i.e., p < 0.05; 95% C.I), are reported in black dots. The correlations are investigated against the residual of COVID-19 cases with a time-shift of 15 days (i.e., with a time-lag of -14 days) to take into consideration the incubation period.

Fig 7 (top panel) highlights that statistically significant inverse correlations (i.e. with p < 0.01; 99% C.I) are clearly found between COVID-19 cases and surface air temperatures over most of the Maghreb countries (Tunisia, Morocco and Libya) and in all of the Gulf of Guinea countries except Togo, while significant positive correlations (i.e. with p < 0.01; 99% C.I) between COVID-19 cases and surface air temperature are found in the Sahel, especially over the central part including Niger and Mali. Despite positive correlations between COVID-19 cases and surface air temperature were found in some specific Maghreb countries such as Egypt and Algeria mainly due to the dry hot weather associated with higher temperatures exceeding 38°C in Egypt and Algeria’s southern regions located in the north and central Sahara as shown in Figs 5A and 5C. Nevertheless, on average, statistically significant inverse correlations between COVID-19 cases and temperature are highlight over the Maghreb and the Gulf of Guinea, whereas positive correlations are found in the Sahel.

Considering the correlations with humidity and water vapor parameters, the Fig 7 (middle and bottom panels) display significant and negative values over all the Maghreb countries and positive values is most countries over the Gulf of Guinea. Over the Sahel, the significant and high positive correlations (τ=0.6, p < 0.01; 99% C.I) are observed over Senegal where the highest transmission of the COVID-19 pandemic are reported (Fig 3B), while negative and non-significant correlations can be observed in Burkina Faso, Mali and Niger. Comparing S1 Fig (i.e., correlation values calculated with Spearman rank test) and results shown in Fig 7 (i.e., Kendall rank test versus Spearman rank test), our results are even more robust because all correlations have the same sign. The magnitude of correlations is however lower in the Kendall test mainly because of its high insensitivity to error and discrepancies in datasets compared to Spearman rank test.

On the other hand, given the average, our results highlight that the COVID-19 pandemic transmission is differently affected across the three bioclimatic regions:

i. cold and dry environmental conditions over the Maghreb; these results imply that over the Maghreb countries, the COVID-19 transmission is strongly influenced by a decrease of temperature that connects to a decrease of humidity and water vapor content. These results confirm previous findings [11,12,12,17,18,20–22,38,51,52], that virus transmission is enhanced by cold and dry climates conditions and may partly explain the high number of COVID-19 pandemic transmission observed in autumn over the Maghreb countries (Fig 4A, Fig 6B and S2A Fig).
ii. warm and humid conditions over the Sahel; these results highlight that over the Sahel, the COVID-19 pandemic transmission prefers warm and humid environmental conditions. In other words, COVID-19 is strongly influenced by an increase in temperature associated with an increase in humidity and water vapor. Such findings contrast with the results found over the Maghreb countries and other studies suggesting that summer weather could reduce COVID-19 transmission [53] but confirm some previous findings (e.g. [54]), and may partly explain the high number of COVID-19 pandemic transmission (i.e. the peak) experienced in most countries during the period May-August (Figs 4B and S2B Fig).
iii. relatively cold and humid conditions over the Gulf of Guinea; these results highlight that the COVID-19 transmission over the Gulf of Guinea countries is enhanced by a decrease of temperature associated with an increase of humidity and water vapor. In other words, COVID-19 transmission is strongly enhanced by cooler and humid climates conditions. Such findings also confirm the previous findings (e.g.,[25,26]) and may explain the observed peak of the COVID-19 cases experienced in the Gulf of Guinea countries during the period May-August (Fig 4C and Fig 6E and S2C Fig).

## Conclusions

An extension of the analysis available in literature for the quantification of the relationship between climatic variables and COVID-19 pandemic transmission in North and West Africa is presented in this paper. The contributions of climate conditions on the COVID-19 transmission have been studied in 16 most highly populated West and North African countries divided into three bioclimatic regions (i.e., the Maghreb, the Sahel and the Gulf of Guinea) based on Köppen bioclimatic classification [32]. The correlation between the basic meteorological variables and virus transmission over 275 days was investigated from March 1, 2020 to November 30, 2020 at each of the considered country and at regional level. To assess the correlation, differently from other studies [29–31], here, we considered as a reliable variable the residuals of the daily new positive cases with respect to the robust and non-parametric Median-Based Linear Model (MBLM) defined in [49]. Further, when working on the residuals, makes our analysis independent on the analyzed time period. Also, we have taken into consideration the incubation period, i.e., the correlations were investigated against the residual of COVID-19 cases with a time-shift of 15 days, -i.e., meteorological data from 15 February 2020 to 15 November 2020 was used in the analysis. The strongest finding of this paper is that the COVID-19 pandemic transmission is differently affected across the three bioclimatic regions. First of all, over the Maghreb countries, our results highlight that the COVID-19 transmission is strongly enhanced by a decrease of temperature associated with a decrease of humidity and water vapor, i.e., cold and dry environmental conditions. Such findings are consistent with previous studies available in the literature [11,12,17,18,20–22,30,38,52,55] supporting the hypothesis that COVID-19 transmission prefers cold and dry weather conditions, while more robust because they are often limited on a single country or city and/or considering a short period of COVID-19 reported data. Over the Sahel, our results show that the COVID-19 pandemic transmission prefers warm and humid climate conditions. In other words, the virus transmission is strongly influenced by an increase in temperature associated with an increase in humidity and water vapor. Such findings contrast with the studies suggesting that summer weather could reduce COVID-19 transmission (e.g.[53]) but are consistent with some previous studies available in the literature (e.g. [54]) and [23] (for a review), and may partly explain the high number of COVID-19 cases experienced in most of Sahelian countries during the period May-August (Fig 4B and S2B Fig). Finally, over the Gulf of Guinea, cold and humid conditions are found to enhance the COVID-19 transmission. In other words, COVID-19 transmission is strongly enhanced by a decrease of temperature associated with an increase of humidity and water vapor. Such findings also confirm the previous findings available in the literature (e.g. [25,26]) and may related to the observed peak of number cases experienced in most of the Gulf of Guinea countries during the period May-August (Fig 4C and Fig 6E and S2C Fig).

Another important aspect of this paper is that this is, to our knowledge, the first work based in three bioclimatic regions (i.e., the Maghreb, the Sahel and the Gulf of Guinea) using COVID-19 datasets reported over 275 days (i.e., 09 months) and investigating the correlation against the residual of COVID-19 cases with a time-shift of 15 days, i.e., taking into consideration the incubation period. The results from this analysis suggest that further studies are needed to investigate why in three considered bioclimatic regions, the virus transmission is differently affected by the climate conditions. These results can promote further studies in other bioclimatic regions testing also the effects of air-pollution related variables, such as the concentration of the Particulate Matter with an aerodynamic diameter less than 2.5 micron (PM2.5), the Nitrogen Dioxide (NO_2_) and Carbon Monoxide. Therefore, this work contributes to increasing attention and encourages further studies to improve our understanding of the underlying factors that control the relationship between climate conditions and COVID-19.

## Data Availability

All data are fully available without restriction
The data underlying the results presented in the study are available from: https://www.ecdc.europa.eu/en/publications-data/download-todays-data-geographicdistribution-covid-19-cases-worldwide (for the COVID-19 Data)
http://iridl.ldeo.columbia.edu/SOURCES/.NOAA/.NCEP-NCAR/.CDAS1/.DAILY/.Intrinsic/.PressureLevel/ (for the Climate Data)

## Acknowledgments

We thank the Ministère de l’Enseignement supérieur de la Recherche et de l’Innovation (MESRI) Sénégal du Sénégal; whose grant awarded the achievement of this study. We would also like to thank the different institutions from which the authors of this work are affiliated.

## Author contributions

A.T.G and W.T. established our consortium and initiated the project. I.D assisted by S.S, H.S, P.F, D.D and M.D performed the experimental activities, analyzed the data, and drafted and wrote the initial manuscript. A.T.G and W.T. designed and supervised the study and performed discussion related to COVID-19 context. S.S contributed significantly to the data analysis, interpretations and the discussions of the results and improved the draft. All authors discussed the results and helped to improve the manuscript. All authors agree on the final version of the manuscript.

## Data Availability

The data underlying the results presented in the study are available from: https://www.ecdc.europa.eu/en/publications-data/download-todays-data-geographicdistribution-covid-19-cases-worldwide (for the COVID-19 Data)

http://iridl.ldeo.columbia.edu/SOURCES/.NOAA/.NCEP-NCAR/.CDAS1/.DAILY/.Intrinsic/.PressureLevel/ (for the Climate Data)

