## Supplementary material for "Potential contribution of climate conditions on COVID-19 pandemic transmission over West and North African countries": S1_Figure, S2_figure and S1_Table: S1_Figure.pdf

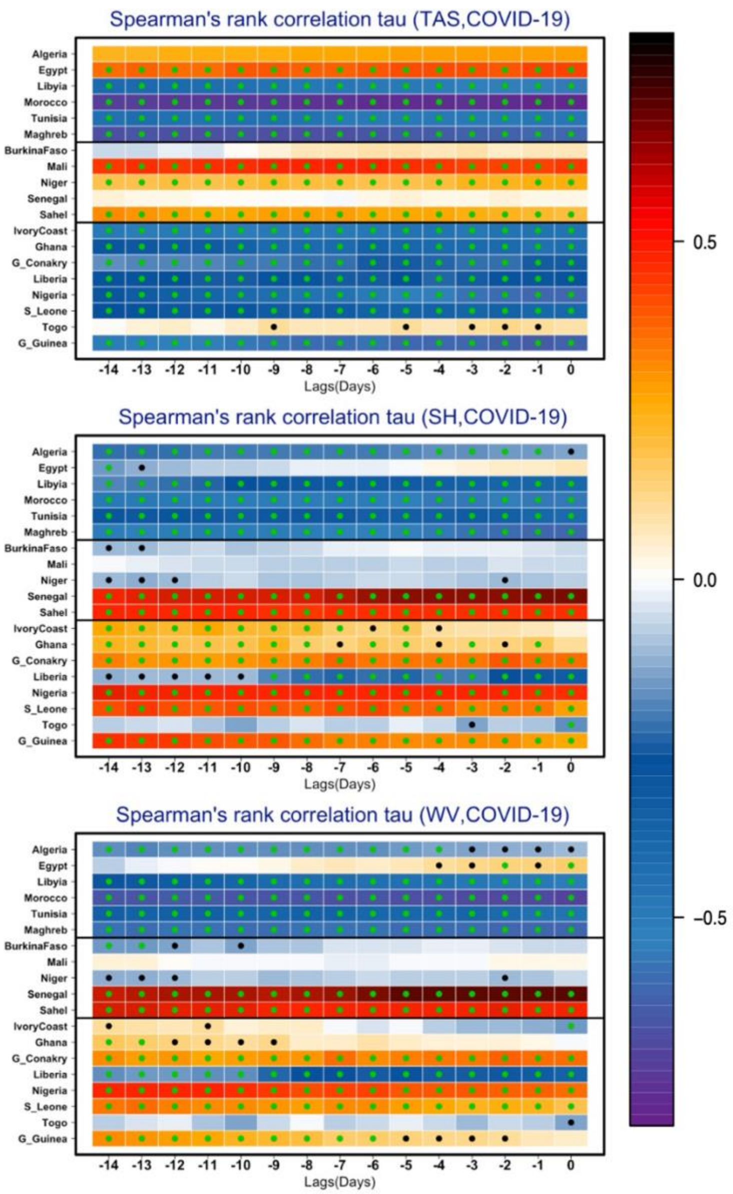


**S1 Fig. Spearman non-linear rank correlation (τ) test between the detrended anomaly of the meteorological variables and the residuals of COVID-19 cases over the selected Northern and Western African countries, as well as area averages over the three bioclimatic regions: the Maghreb, the Sahel and Gulf of Guinea.** Statistically significant correlation with 99% confidence intervals, that is, correlation that passed the Spearman rank tests at 0.01 level (i.e., $p < 0.01$; 99% C.I), is reported in green dot, while correlation that passed the Spearman rank tests at 0.05 level (i.e., $p < 0.05$; 95% C.I), is reported in black dot. The correlations are investigated against the residual of COVID-19 cases with a time-shift of 15 days to take into consideration the incubation period.
