## Supplementary material for "Potential contribution of climate conditions on COVID-19 pandemic transmission over West and North African countries": S1_Figure, S2_figure and S1_Table: S1_Table.pdf

**S1 Table. Descriptive statistical analyses of meteorological variables (February. 15 to November 15, 2020; N = 275) over the 16 selected Northern and Western African countries, as well as area averages over the three bioclimatic regions: the Maghreb, the Sahel and Gulf of Guinea**.

| Climate Parameters | Temperature (Mean ± SD) | Specific Humidity (Mean ± SD) | Water Vapor (Mean ± SD) | Daily Cases (Mean ± SD) |
|---|---|---|---|---|
| **Algeria** | 28.45± 7.08 | 6.20±1.25 | 11.027±2.55 | 298.98±259.33 |
| **Egypt** | 24.59 ± 6.07 | 8.41±2.35 | 13.83±4.20 | 420.14±487.73 |
| **Libya** | 25.67 ± 6.77 | 6.77±1.83 | 11.45±3.41 | 299.74± 379.68 |
| **Morocco** | 27.75±6.55 | 7.10±1.44 | 12.67±3.10 | 1286.55± 1620.26 |
| **Tunisia** | 22.84±6.55 | 9.06±2.52 | 14.65±4.54 | 350.00±744.50 |
| **MAGRHEB** | 25.86±6.24 | 7.51±1.60 | 13.18±3.40 | 531.08±503.38 |
| **Burkina Faso** | 29.64±3.23 | 14.77±5.71 | 23.55±8.77 | 10.37±15.11 |
| **Mali** | 31.0±3.26 | 11.60±5.32 | 19.32±8.814 | 16.94±21.7 |
| **Niger** | 30.49±4.03 | 11.38±5.70 | 18.89±9.33 | 5.51±10.86 |
| **Senegal** | 28.63±1.94 | 12.86±4.95 | 20.53±7.544 | 58.45±49.82 |
| **SAHEL** | 29.96±2.76 | 12.65±5.23 | 20.72±8.37 | 22.82±13.67 |
| **Ivory Coast** | 27.022±2.09 | 18.15±2.64 | 28.08±3.75 | 77.93±101.34 |
| **Ghana** | 27.43±2.52 | 17.45±3.34 | 27.179±4.84 | 187.52±264.26 |
| **Guinea Conakry** | 27.92±2.42 | 17.04±3.37 | 26.59±4.85 | 47.41±42.87 |
| **Liberia** | 26.60±1.72 | 19.27±1.68 | 29.53±2.48 | 5.80±8.28 |
| **Nigeria** | 28.05±2.47 | 15.98±5.11 | 25.21±7.83 | 245.13±2.47 |
| **Sierra Leone** | 27.20±1.96 | 18.64±2.30 | 28.74±3.25 | 8.76±10.94 |
| **Togo** | 27.49±2.70 | 17.16±3.88 | 26.67±5.67 | 10.77±9.67 |
| **GULF OF GUINEA** | 27.39±2.16 | 17.67±2.89 | 27.50±4.21 | 83.33±72.59 |
