## Supplementary material for "Potential contribution of climate conditions on COVID-19 pandemic transmission over West and North African countries": S1_Figure, S2_figure and S1_Table: S2_Figure.pdf

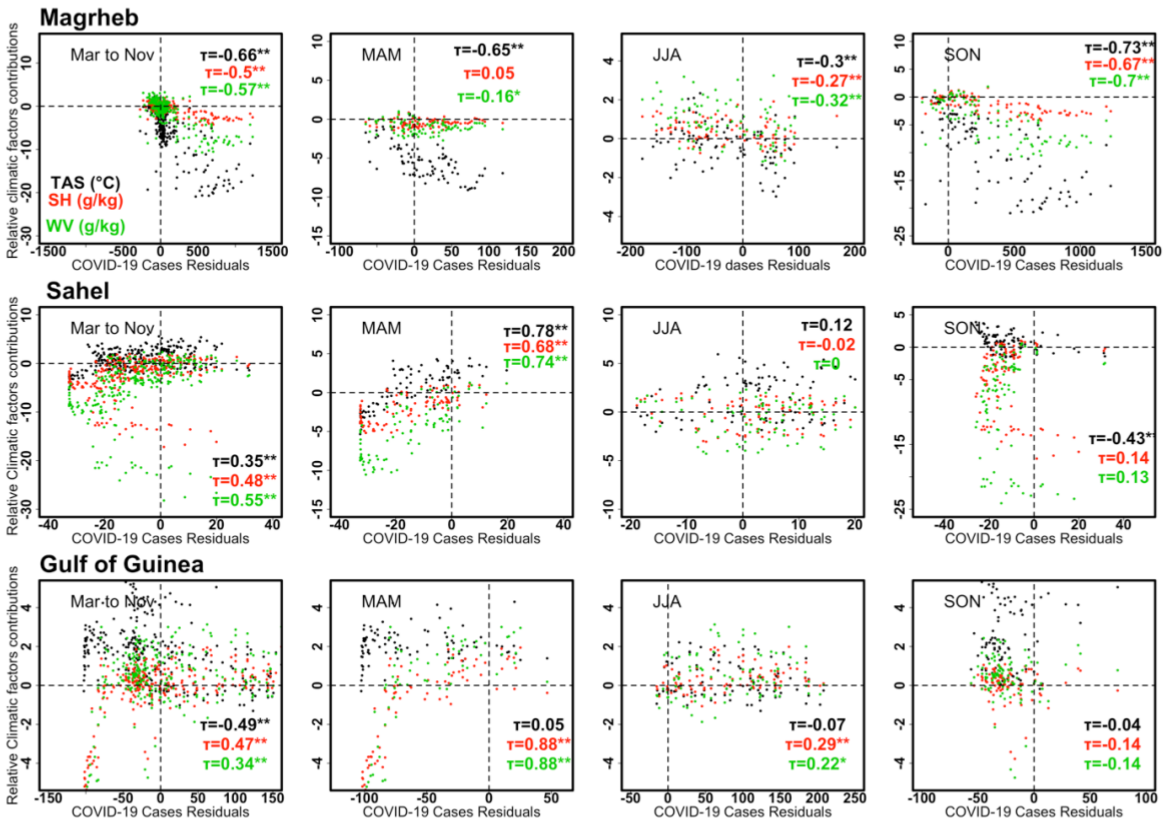


**S2 Fig. Seasonal effects of the relative climate factors contributions on the COVID-19 pandemic transmission.** The relation between COVID-19 pandemic transmission (x axis) versus (1) Temperature (TAS °C) in black, (2) Specific Humidity (SH, g/kg) in red, (3) Water Vapor (WV, g/kg) in green, is drawn (y-axis). Correlations are calculated for each contribution using Kendall non-linear rank (τ) test between the detrended anomaly of the meteorological variables and the residuals of COVID-19 cases over the three bioclimatic regions. Two star symbols (**) are added when the correlation is significant at 99% confidence intervals (i.e., $p < 0.01$; 99% C.I). A star symbol (*) is added when the correlation is significant at 95% confidence intervals (i.e., $p < 0.05$; 95% C.I). The correlation is investigated against the residual of COVID-19 cases with a time-shift of 15 days to take into consideration the incubation period.
